# Determinants of Domestic Violence Against Women in Cambodia: How Digital Access, Media Exposure, Motorcycle Ownership, and Partners’ Alcohol Use Matter

**DOI:** 10.1101/2025.08.28.25334636

**Authors:** Samnang Um, Sopheap Suong, Chantrea Sieng, Sovandara Heng, Grace Marie Ku, Sothy Heng

**Author notes:** Address: Lot#: 80, 289 Samdach Penn Nouth St. (289), Phnom Penh.

## Abstract

Domestic violence against women remains a public health and socio-economic burden in Cambodia, with slow declines over the past two decades. This study examined how digital access, media exposure, motorcycle ownership, and partners’ alcohol use are associated with violence types: physical, sexual, emotional, and intimate partner violence (IPV), while controlling for socio-demographic factors. A cross-sectional study utilized secondary data, analyzing 5,780 weighted women aged 15-49 in the 2021-2022 Cambodia Demographic and Health Survey. IPV—defined as sexual, physical, or emotional violence in the previous 12 months—was regressed on mobile phone ownership, internet use, media exposure, motorcycle ownership, and partners’ alcohol use using survey-adjusted multivariable logistic models. Overall, 13.2% reported experiencing intimate partner violence (IPV) in the past year, with emotional violence at 12.2%, physical violence at 4.4%, and sexual violence at 1.9%. Women’s smartphone ownership was associated with lower odds of emotional violence (AOR = 0.7; 95 % CI 0.5–0.9) and IPV (AOR = 0.7, 0.5–1.0) relative to no-phone users. Conversely, low-frequency internet use predicted higher odds of emotional violence (AOR=1.7, 1.1–2.7) and IPV (AOR = 1.6, 1.1–2.5). Partner alcohol use is a strong risk factor for all violence types: sexual (AOR = 3.5; 95% CI (1.1-11.4), physical (AOR = 5.6; 95% CI (2.8-11.5), emotional (AOR = 3.1; 95% CI (2.2-4.4), and IPV (AOR=3.0; 95% CI (2.1-4.1). Inversely, women in rich households had significantly lower odds of physical (AOR = 0.4; 95% CI (0.3–0.7) and IPV (AOR = 0.6; 95% CI (0.5–0.8). Digital inclusion appears to have a dual role—smartphones may protect women from physical, sexual, emotional, and intimate partner violence (IPV), yet sporadic internet access could heighten the risk, possibly by triggering partner suspicion. Policies that expand safe, private digital access while tackling alcohol misuse and poverty may reduce domestic violence in Cambodia.

## Introduction

Domestic violence against women, particularly intimate partner violence (IPV), remains a serious public health and women’s human rights concern (1). Estimates by the World Health Organization (WHO) indicate that nearly 1 in 3 (30%) of women worldwide have experienced either physical and/or sexual intimate partner violence in their lifetime (1). This issue is particularly more observed in low- and middle-income countries, where gender inequality, poverty, and limited access to resources increase women’s vulnerability (2–4).

In Southeast Asia, the estimated rate of IPV stands at approximately 33% of women(1). In Cambodia, despite policy and legal reforms, domestic violence remains a pressing issue (5, 6). The 2021–2022 Cambodia Demographic and Health Survey (CDHS) showed that 22% of women aged 15–49 had experienced emotional, physical, or sexual violence by a husband or partner at some point, and 8% of survey respondents reported such experiences in the 12 months before the survey (7). More specifically, 17% of women reported physical violence, 15% emotional violence, and 4% sexual violence from their intimate partners (7). These forms of abuse often overlap, and many women endure more than one type of violence (1–4, 6–8). The long-term impacts are profound. Beyond physical injuries, survivors face higher risks of depression, anxiety, reproductive health problems, and social isolation. Domestic violence also restricts women’s economic participation and reinforces cycles of poverty and dependence. In Cambodia, data suggest a slow decline in IPV prevalence. For instance, physical violence reported by ever-partnered women dropped from 23% in 2005 to 20% in 2014 and 17% in 2021– 22, indicating gradual progress (7–9). The persistence of such violence—especially in rural areas—signals the need for more targeted, structural interventions (7–9).

While most research on domestic violence in Cambodia have focused on socio-demographic factors such as age, education level, income, occupation, or alcohol use by partners, emerging evidence points to new areas of influence (9). Access to digital technology, exposure to mass media, and ownership of transportation such as motorcycles are particularly being explored as potential factors that could reduce the risk of violence. However, there is still limited research on these aspects in the Cambodian context.

Digital access—such as mobile phone and internet use—can increase women’s autonomy, improve access to information and services, and provide discreet channels for help-seeking. Studies in India and Africa show a positive link between mobile phone ownership and reduced IPV risk, likely due to increased social connectedness and reduced isolation (10–14).

Media exposure through radio, television, or newspapers can positively influence social norms by challenging harmful gender stereotypes and encouraging non-violent conflict resolution (15). Research from Bangladesh and Nigeria have shown that women with regular media exposure are less tolerant of domestic violence and more likely to seek help (16–18).

Motorcycle ownership and mobility also play a critical role in addressing domestic violence. In areas with poor transportation infrastructure, owning a motorcycle can enhance a woman’s ability to access services, participate in the workforce, or leave abusive situations. Evidence from rural India and Southeast Asia suggests that women with access to transportation are more empowered and better positioned to seek support or exit violent relationships (19–21).

Cambodia’s National Action Plan to Prevent Violence Against Women 2019-2023 underscores the importance of multisectoral strategies to prevent and respond to gender-based violence, with a specific focus on empowerment through access to information, services, and technology (22). The plan recognizes the transformative role of digital connectivity, media, and mobility in reshaping gender norms and increasing women’s agency. This is consistent with global evidence suggesting that digital inclusion, media literacy, and personal mobility may influence women’s exposure to IPV. For example, mobile phone and internet access can enhance women’s autonomy, improve help-seeking, and reduce isolation. Likewise, media exposure can challenge harmful social norms, and access to transportation such as motorcycles may increase women’s capacity to access services or leave abusive environments.

Despite these promising pathways, few studies in Cambodia have empirically examined the relationship between digital access, media exposure, and mobility on IPV risk. This study seeks to fill that gap by analyzing data from the 2021–2022 CDHS to assess the associations between smartphone and internet access, media exposure, motorcycle ownership, and partners’ alcohol use, and women’s experiences of violence types—while accounting for key sociodemographic and contextual factors. The findings aim to inform the implementation and future directions of national strategies like the National Action Plan to Prevent Violence Against Women and other gender-responsive development efforts.

## Methods

### Ethical statement

The Cambodia National Ethics Committee for Human Health Research (NECHR) approved the data collection tools and procedures for CDHS 2021-2022 for Health Research on May 10, 2021 (Ref # 83 NECHR), and ICF’s Institutional Review Board (IRB) in Rockville, Maryland, USA. Written informed consent was obtained from all participants prior to data collection. For respondents under 18 years of age, consent was obtained from a parent or guardian. This study used de-identified secondary data and was therefore exempt from additional institutional ethical approval.

### Data Source

We used data from the 2021–2022 Cambodia Demographic and Health Survey (CDHS), a nationally representative cross-sectional survey conducted by the National Institute of Statistics and the Ministry of Health, with technical support from ICF International. Data was collected from September 15, 2021, to February 15, 2022. Two-stage stratified cluster sampling was used to collect the samples (7). First, 709 clusters or enumeration areas (EAs) were selected and stratified by urban-rural using probability proportional to cluster size, and second, 25-30 households were selected in each EA using systematic sampling (7). In total, 19,496 women aged 15-49 were interviewed face-to-face, using the survey questionnaire, including socio-demographic characteristics, alcohol drinking, tobacco use, household assets, maternal health-related indicators, and nutritional status (7). The domestic violence module was administered to a subsample of 6,204 eligible women (5,780 weighted), in accordance with WHO ethical guideline (7, 23).

### Study Population

The analysis included 5,780 weighted women aged 15–49 who completed the domestic violence module and had complete data on key variables of interest (7).

### Outcome Variable

The primary outcome was experience of domestic violence, defined as self-reported experience of sexual violence, physical violence, emotional violence, and intimate partner violence (IPV) by a husband or partner in the past 12 months (7).

### Independent Variables

**Main exposure:** Women’s characteristics included mobile phone ownership (categorized as no mobile phone, non-smartphone, and smartphone), internet use frequency (no use, low frequency, and daily or almost daily use), and media exposure measured by the number of media types accessed (none, one, or two or more of newspaper, radio, and television). Additionally, household motorcycle ownership (yes/no). **Covariates**: women’s age groups (<24, 25–34, 35–49 years), marital status (not married, married, widowed/divorced), educational attainment (no education, primary, secondary or above), and employment status (employed or not employed) were considered. Partner characteristics incorporated age groups (<24, 25–34, 35–44, ≥45 years), educational levels (no education, primary, secondary, or above), and alcohol use (yes/no). Household wealth status was categorized into three quintiles (poor, middle, and rich) based on the CDHS reported (7), and place of residence (urban or rural).

### Statistical Analysis

The data was analyzed using STATA v18 (Stata Corp, Texas, 2023) (24). The standard DHS sampling weight and complex survey design were accounted for using the survey package. Descriptive statistics were used to estimate key characteristics of the study population, with results presented as weighted frequencies and percentages.

A separate bivariate analysis using chi-square tests was conducted to explore associations between independent variables (mobile phone ownership, internet use frequency, media exposure, partners’ alcohol use, and other covariates) with outcome variables (sexual violence, physical violence, emotional violence, and IPV). Variables that showed a significant association (p-value ≤ 0.05) in the bivariate chi-square analysis were then included in the multiple logistic regression analysis [21].

Multiple logistic regression models were used to examine the associations between the independent variables (digital access, media exposure, motorcycle ownership, and partner’s alcohol use) and the significant outcome variables, while controlling for covariate variables. Results are reported as adjusted odds ratios (AOR) with 95% confidence intervals (CI).

Multicollinearity of the independent variables (mobile phone ownership, internet use frequency, media exposure, partners’ alcohol use, age, wealth index, education, and place of residence) was evaluated using the variance inflation factor (VIF) for the regression coefficients (25).

The goodness-of-fit of the logistic regression models for different forms of violence types was evaluated using the F-adjusted mean residual goodness-of-fit test.

To evaluate the potential effect modification of statistically significant associations in the adjusted analysis, these were further visualized as predicted probabilities using STATA 18, employing the margins command to estimate and visualize with a marginal plot (26).

## Results

### Descriptive Characteristics of the Study Population

This study analyzed data from 5,780 Cambodian women aged 15-49 years. Most participants (73.7%) reported owning a smartphone, while 17.1% did not own any mobile phone. About half (51.5%) used the internet daily, whereas 38.9% had never used it. More than half of the women (54.4%) reported no regular exposure to newspapers, radio, or television. Motorcycle ownership was common, reported by 87.0% of respondents. Most women were married or living with a partner (87.2%), and 70.5% reported employment. Educational levels varied: 13.7% had no formal schooling, while 42.5% had reached at least secondary school. Looking at household wealth, 44.7% lived in richer households, 35.9% in poorer ones, and 19.5% fell in the middle category. Half (50.1%) of male partners had completed secondary education or higher, while 10.4% had no schooling. Alcohol use among male partners was reported by 82.4% of participants (**Table 1**).

**Table 1.** Descriptive Characteristics of the Study Population (N = 5,780 weighted).

| Variables | Frequency | Percentage (%) |
| --- | --- | --- |
| **Women's Characteristics** |  |  |
| Mobile Phone Ownership |  |  |
| No mobile | 990 | 17.1 |
| Non-smartphone | 531 | 9.2 |
| Smartphone | 4259 | 73.7 |
| Internet Use |  |  |
| No internet use | 2250 | 38.9 |
| Low-frequency use | 556 | 9.6 |
| Daily/Almost daily | 2974 | 51.5 |
| Media exposure (Newspaper, radio, TV) |  |  |
| None | 3146 | 54.4 |
| One | 1820 | 31.5 |
| Two or more | 815 | 14.1 |
| Motorcycle Ownership |  |  |
| No | 750 | 13.0 |
| Yes | 5031 | 87.0 |
| Women's Age |  |  |
| ≤ 24 | 922 | 15.9 |
| 25-34 | 2155 | 37.3 |
| 35-49 | 2703 | 46.8 |
| Marital Status |  |  |
| Not married | 267 | 4.6 |
| Married | 5042 | 87.2 |
| Widowed/Divorced | 472 | 8.2 |
| Educational |  |  |
| No education | 790 | 13.7 |
| Primary | 2535 | 43.9 |
| Secondary or above | 2455 | 42.5 |
| Employment Status |  |  |
| No | 1708 | 29.5 |
| Yes | 4072 | 70.5 |
| **Partner's Characteristics** |  |  |
| Partner's Age Group |  |  |
| ≤ 24 | 343 | 6.8 |
| 25-34 | 1712 | 34.0 |
| 35-44 | 2002 | 39.7 |
| 45+ | 984 | 19.5 |
| Partner's Education |  |  |
| No education | 526 | 10.4 |
| Primary | 1992 | 39.5 |
| Secondary or above | 2524 | 50.1 |
| Partner's Alcohol Use |  |  |
| No | 1018 | 17.6 |
| Yes | 4763 | 82.4 |
| **Household Characteristics** |  |  |
| Wealth Index |  |  |
| Poor | 2073 | 35.9 |
| Middle | 1125 | 19.5 |
| Rich | 2583 | 44.7 |
| Place of Residence |  |  |
| Urban | 2379 | 41.1 |
| Rural | 3402 | 58.9 |

*Notes: Survey weights are applied to obtain weighted percentages.*

### Prevalence of Sexual, Physical, Emotional, and Intimate Partner Violence among Women

Over 13.2% reported experiencing intimate partner violence (IPV) in the past year, whether physical, emotional, or sexual. Emotional violence emerged as the most common type, affecting 12.2% of women. A total of 4.4% reported physical violence, while sexual violence was the least reported, with 1.9% of women disclosing such experiences (**Table 2**).

**Table 2.** Prevalence of Sexual, Physical, Emotional, and Intimate Partner Violence Among Women (N = 5,780 weighted).

| Type of Violence | No. of Women | Percentage (%) |
| --- | --- | --- |
| Sexual Violence |  |  |
| No | 5673 | 98.1 |
| Yes | 107 | 1.9 |
| Physical Violence |  |  |
| No | 5525 | 95.6 |
| Yes | 255 | 4.4 |
| Emotional Violence |  |  |
| No | 5076 | 87.8 |
| Yes | 704 | 12.2 |
| Intimate Partner Violence (IPV) |  |  |
| None | 5019 | 86.8 |
| One or more | 761 | 13.2 |

— **Notes:** Survey weights are applied to obtain weighted percentages.

### Factors Associated with sexual violence, physical violence, emotional violence, and intimate partner violence (IPV) Among Cambodian Women Aged 15–49 Years in Chi-Square Analysis

**Table 3** illustrates that the prevalence of sexual violence, physical violence, emotional violence, and intimate partner violence (IPV) among ever-partnered women aged 15–49 in Cambodia varied across individual, partner, household, and contextual factors.

**Table 3.** Prevalence of Sexual Violence, Physical Violence, Emotional Violence, and Intimate Partner Violence Among Women by Background Characteristics using Chi-Square test, Cambodia DHS 2021–2022, (N = 5,780 weighted).

| Variables | Sexual<br>(N=107) |  | P | Physical<br>(N=225) |  | P | Emotional<br>(N=704) |  | P | IPV (N=761) |  | P |
| --- | --- | --- | --- | --- | --- | --- | --- | --- | --- | --- | --- | --- |
|  | Freq. | % |  | Freq. | % |  | Freq. | % |  | Freq. | % |  |
| Women's Characteristics |  |  |  |  |  |  |  |  |  |  |  |  |
| Mobile Phone Ownership |  |  |  |  |  |  |  |  |  |  |  |  |
| No mobile | 41 | 4.1 | <0.001 | 75 | 7.6 | <0.001 | 175 | 17.7 | <0.001 | 188 | 19.0 | <0.001 |
| Non-smartphone | 13 | 2.4 |  | 28 | 5.3 |  | 91 | 17.1 |  | 95 | 17.9 |  |
| Smartphone | 53 | 1.2 |  | 152 | 3.6 |  | 438 | 10.3 |  | 478 | 11.2 |  |
| Internet Use |  |  |  |  |  |  |  |  |  |  |  |  |
| No internet use | 65 | 2.9 | <0.001 | 136 | 6.0 | <0.001 | 330 | 14.7 | <0.001 | 356 | 15.8 | <0.001 |
| Low-frequency use | 7 | 1.3 |  | 38 | 6.8 |  | 96 | 17.3 |  | 103 | 18.5 |  |
| Daily/Almost daily | 35 | 1.2 |  | 81 | 2.7 |  | 278 | 9.3 |  | 303 | 10.2 |  |
| Media exposure (Newspaper, radio, TV) |  |  |  |  |  |  |  |  |  |  |  |  |
| None | 59 | 1.9 | 0.990 | 152 | 4.8 | 0.378 | 394 | 12.5 | 0.291 | 430 | 13.7 | 0.302 |
| One | 33 | 1.8 |  | 73 | 4.0 |  | 230 | 12.6 |  | 244 | 13.4 |  |
| Two or more | 15 | 1.8 |  | 30 | 3.7 |  | 80 | 9.8 |  | 88 | 10.8 |  |
| Motorcycle Ownership |  |  |  |  |  |  |  |  |  |  |  |  |
| No | 21 | 2.8 | 0.082 | 43 | 5.7 | 0.109 | 108 | 14.4 | 0.087 | 116 | 15.5 | 0.083 |
| Yes | 86 | 1.7 |  | 212 | 4.2 |  | 597 | 11.9 |  | 645 | 12.8 |  |
|  | Freq. | % |  | Freq. | % |  | Freq. | % |  | Freq. | % |  |
| Women's Age |  |  |  |  |  |  |  |  |  |  |  |  |
| < 24 | 7 | 0.8 | 0.004 | 24 | 2.6 | 0.007 | 76 | 8.2 | <0.001 | 85 | 9.2 | <0.001 |
| 25-34 | 34 | 1.6 |  | 81 | 3.8 |  | 226 | 10.5 |  | 248 | 11.5 |  |
| 35-49 | 67 | 2.5 |  | 150 | 5.5 |  | 403 | 14.9 |  | 428 | 15.8 |  |
| Marital Status |  |  |  |  |  |  |  |  |  |  |  |  |
| Not married | 0 | 0.0 | 0.190 | 1 | 0.4 | 0.004 | 4 | 1.5 | <0.001 | 4 | 1.5 | <0.001 |
| Married | 95 | 1.9 |  | 220 | 4.4 |  | 640 | 12.7 |  | 690 | 13.7 |  |
| Widowed/Divorced | 13 | 2.8 |  | 34 | 7.2 |  | 60 | 12.7 |  | 67 | 14.2 |  |
| Educational |  |  |  |  |  |  |  |  |  |  |  |  |
| No education | 30 | 3.8 | 0.002 | 62 | 7.8 | <0.001 | 151 | 19.1 | <0.001 | 156 | 19.7 | <0.001 |
| Primary | 47 | 1.9 |  | 139 | 5.5 |  | 362 | 14.3 |  | 392 | 15.5 |  |
| Secondary or above | 30 | 1.2 |  | 54 | 2.2 |  | 192 | 7.8 |  | 214 | 8.7 |  |
| Employment Status |  |  |  |  |  |  |  |  |  |  |  |  |
| No | 32 | 1.9 | 0.973 | 70 | 4.1 | 0.501 | 231 | 13.5 | 0.090 | 245 | 14.3 | 0.153 |
| Yes | 76 | 1.9 |  | 185 | 4.5 |  | 473 | 11.6 |  | 516 | 12.7 |  |
| **Partner's Characteristics** |  |  |  |  |  |  |  |  |  |  |  |  |
| Partner's Age |  |  |  |  |  |  |  |  |  |  |  |  |
| ≤ 24 | 5 | 1.5 | 0.035 | 13 | 3.8 | 0.026 | 34 | 9.9 | <0.001 | 38 | 11.1 | 0.001 |
| 25-34 | 17 | 1.0 |  | 55 | 3.2 |  | 166 | 9.7 |  | 183 | 10.7 |  |
| 35-44 | 46 | 2.3 |  | 93 | 4.6 |  | 291 | 14.5 |  | 313 | 15.6 |  |
| 45+ | 27 | 2.7 |  | 59 | 6.0 |  | 149 | 15.1 |  | 156 | 15.8 |  |
| Partner's Education |  |  |  |  |  |  |  |  |  |  |  |  |
| No education | 18 | 3.4 | 0.045 | 45 | 8.5 | <0.001 | 120 | 22.8 | <0.001 | 129 | 24.5 | <0.001 |
| Primary | 41 | 2.1 |  | 97 | 4.9 |  | 275 | 13.8 |  | 296 | 14.9 |  |
| Secondary or above | 35 | 1.4 |  | 79 | 3.1 |  | 246 | 9.7 |  | 266 | 10.5 |  |
| Partner's Alcohol Use |  |  |  |  |  |  |  |  |  |  |  |  |
| No | 7 | 0.7 | 0.023 | 15 | 1.5 | <0.001 | 54 | 5.3 | <0.001 | 59 | 5.8 | <0.001 |
| Yes | 101 | 2.1 |  | 240 | 5.0 |  | 650 | 13.6 |  | 702 | 14.7 |  |
| **Household Characteristics** |  |  |  |  |  |  |  |  |  |  |  |  |
| Wealth Index |  |  |  |  |  |  |  |  |  |  |  |  |
| Poor | 57 | 2.8 | 0.012 | 150 | 7.2 | <0.001 | 354 | 17.1 | <0.001 | 384 | 18.5 | <0.001 |
| Middle | 20 | 1.8 |  | 49 | 4.4 |  | 134 | 11.9 |  | 151 | 13.4 |  |
| Rich | 30 | 1.2 |  | 56 | 2.2 |  | 216 | 8.4 |  | 226 | 8.8 |  |
| Place of Residence |  |  |  |  |  |  |  |  |  |  |  |  |
| Urban | 35 | 1.5 | 0.212 | 69 | 2.9 | <0.001 | 211 | 8.9 | <0.001 | 225 | 9.5 | <0.001 |
| Rural | 72 | 2.1 |  | 186 | 5.5 |  | 493 | 14.5 |  | 536 | 15.8 |  |

**Notes:** Survey weights are applied to obtain weighted percentages. \* IPV: Intimate Partner Violence (includes any of the three forms: sexual, physical, or emotional). \*P: P-value

### Sexual Violence

Sexual violence was more commonly reported by women without access to mobile phones (4.1%) or internet (3.4%), compared to those with smartphones (2.2%) or daily internet use (1.8%) (p < 0.001 for both). Women with no education (5.2%) and those whose partners had no formal education (7.2%) were also more likely to report sexual violence (p < 0.001). Higher prevalence was observed among married or formerly married women (3.4–3.6%) compared to unmarried women (0.4%) (p < 0.001). Partner alcohol use was a significant factor: women whose partners drank alcohol reported more sexual violence (3.5%) than those whose partners did not (1.0%) (p < 0.001).

### Physical Violence

Physical violence was most prevalent among women aged 35–49 (10.7%) and those from poor households (11.8%) (p < 0.001). Women with no education (13.6%) and those whose partners lacked formal education (18.3%) reported higher levels of physical violence (p < 0.001). Living in rural areas (10.1%) and having a partner who used alcohol (10.6%) were also associated with greater physical violence (p < 0.001). Conversely, lower rates were reported among women with higher education (5.3%) and those with daily internet access (4.8%).

### Emotional Violence

Emotional violence was more frequent among women aged 35–49 (9.4%) and those who were formerly married (13.3%) or currently married (7.3%) (p < 0.001). Lack of education in women (9.9%) and their partners (13.1%), rural residence (7.8%), and partner alcohol use (8.3%) were all linked to higher emotional violence (p < 0.001). Women with smartphones (5.7%) and daily internet access (5.3%) reported less emotional violence.

### Intimate Partner Violence (IPV)

Overall, IPV prevalence was significantly higher among women with no phone (19.0%), no internet access (15.8%), and those living in rural areas (15.8%) (p < 0.001 for all). IPV increased with age, from 9.2% in women under 24 to 15.8% in those aged 35–49 (p < 0.001). Educational attainment showed a protective effect: women with secondary or higher education experienced lower IPV (8.7%) than those with no education (19.7%) (p < 0.001). Women whose partners had no education (24.5%) or drank alcohol (14.7%) were at greater risk. IPV was also highest among women from poor households (18.5%) compared to rich households (8.8%) (p < 0.001).

### Multicollinearity

To assess multicollinearity among the independent variables included in the regression models for sexual, physical, and emotional health outcomes, the Variance Inflation Factor (VIF) was computed (**S1 Table to S4 Table**). Across all three models, the VIF values remained below the conventional threshold of 5, indicating no evidence of severe multicollinearity. Specifically, the highest VIF values were observed for age (VIF = 2.04) and partner’s age (VIF = 2.00), while all other variables, including internet use, mobile phone ownership, media exposure, use of motor transportation, educational attainment of women and partners, alcohol use, household wealth, and residence, exhibited VIFs ranging from 1.01 to 1.72. The mean VIF across all models was consistently 1.48, suggesting a low overall collinearity structure. These results confirm that the multivariate regression estimates are unlikely to be distorted by multicollinearity among the predictors.

### Goodness-of-fit

The goodness-of-fit of the logistic regression models for different forms of violence types was evaluated using the F-adjusted mean residual goodness-of-fit test. The model for sexual violence demonstrated a good fit (F(9,652) = 0.63, p = 0.7720), and the emotional violence model also showed an acceptable fit (F(9,652) = 1.57, p = 0.1247). However, the model for physical violence suggested a possible misfit (F(9,652) = 2.07, p = 0.0305). Finally, the IPV model indicated an acceptable fit (F(9,652) = 1.42, p = 0.1760).

### Adjusted Odds Ratio of Digital Access, Media Exposure, and Motorcycle Ownership with Sexual Violence, Physical Violence, Emotional Violence, and Intimate Partner Violence (IPV) Among Women

The adjusted odds ratios (AOR) presented in **Table 4** examine the association between digital access, media exposure, motorcycle ownership, Sexual Violence, Physical Violence, Emotional Violence, and Intimate Partner Violence (IPV) among women in Cambodia after controlling for other socio-demographic factors.

**Table 4.** Adjusted Odds Ratios and 95% Confidence Intervals for the Association of Digital Access, Media Exposure, and Motorcycle Ownership with Sexual Violence, Physical Violence, Emotional Violence, and Intimate Partner Violence (IPV) Among Women in Cambodia, CDHS 2021-2022.

| Variables | Sexual Violence<br>N = 5,780 | Physical<br>Violence<br>N = 5,780 | Emotional<br>Violence<br>N = 5,780 | Any IPV<br>N = 5,780 |
| --- | --- | --- | --- | --- |
|  | AOR (95% CI) | AOR (95% CI) | AOR (95% CI) | AOR (95% CI) |
| **Women's Characteristics** |  |  |  |  |
| Mobile Phone Ownership |  |  |  |  |
| No mobile | Ref. | Ref. | Ref. | Ref. |
| Non-smartphone | 0.6 (0.2–1.4) | 0.7 (0.4–1.1) | 0.9 (0.6–1.3) | 0.9 (0.6–1.3) |
| Smartphone | 0.4 (0.2–1.1) | 0.9 (0.5–1.4) | 0.7(0.5–0.9)* | **0.7(0.5–1.0)\*** |
| Internet Use |  |  |  |  |
| No internet use | Ref. | Ref. | Ref. | Ref. |
| Low-frequency use | 0.6 (0.2–1.9) | 1.2 (0.7–2.0) | 1.7(1.1–2.7)* | 1.6(1.1–2.5)* |
| Daily/Almost daily | 1.0 (0.4–2.3) | 0.7 (0.4–1.2) | 1.2 (0.9–1.7) | 1.2 (0.9–1.6) |
| Media exposure (Newspaper, radio, TV) |  |  |  |  |
| None | Ref. | Ref. | Ref. | Ref. |
| One | 1.1 (0.6–2.2) | 1.0 (0.7–1.4) | 1.2 (0.9–1.5) | 1.2 (0.9–1.5) |
| ≥ Two | 1.1 (0.5–2.2) | 1.1 (0.6–2.0) | 1.0 (0.7–1.5) | 1.0 (0.7–1.5) |
| Motorcycle Ownership |  |  |  |  |
| No | Ref. | Ref. | Ref. | Ref. |
| Yes | 0.8 (0.4–1.7) | 1.1 (0.7–1.7) | 1.1 (0.8–1.5) | 1.1 (0.8–1.5) |
| Women's Age |  |  |  |  |
| 15-24 | Ref. | Ref. | Ref. | Ref. |
| 25-34 | 1.5 (0.5–4.6) | 1.9 (1.0–3.8) | 1.0 (0.7–1.5) | 1.1 (0.7–1.5) |
| 35-49 | 1.7 (0.6–5.5) | 2.1 (1.0–4.5) | 1.2 (0.8–1.8) | 1.2 (0.8–1.8) |
| Woman's Education |  |  |  |  |
| No education | Ref. | Ref. | Ref. | Ref. |
| Primary | 0.7 (0.3–1.4) | 0.8 (0.6–1.3) | 0.9 (0.7–1.2) | 1.0 (0.7–1.3) |
| Secondary or above | 0.7 (0.2–1.9) | 0.5(0.3–0.9)* | 0.7(0.5–1.0)* | 0.7 (0.5–1.1) |
| Employment Status |  |  |  |  |
| No | Ref. | Ref. | Ref. | Ref. |
| Yes | 1.3 (0.7–2.5) | 1.2 (0.9–1.7) | 0.9 (0.7–1.1) | 0.9 (0.8–1.2) |
| **Partner's Characteristics** |  |  |  |  |

| Variables | Sexual Violence<br>N = 5,780 | Physical Violence<br>N = 5,780 | Emotional Violence<br>N = 5,780 | Any IPV<br>N = 5,780 |
| --- | --- | --- | --- | --- |
|  | AOR (95% CI) | AOR (95% CI) | AOR (95% CI) | AOR (95% CI) |
| Partner's Age Group |  |  |  |  |
| ≤ 24 | Ref. | Ref. | Ref. | Ref. |
| 25-34 | 0.6 (0.2–2.2) | 0.6 (0.3–1.3) | 0.9 (0.6–1.5) | 0.9 (0.6–1.5) |
| 35-44 | 1.0 (0.2–4.2) | 0.7 (0.3–1.5) | 1.3 (0.7–2.3) | 1.3 (0.7–2.2) |
| 45+ | 1.2 (0.3–4.7) | 0.9 (0.3–2.3) | 1.4 (0.7–2.5) | 1.3 (0.7–2.4) |
| Partner's Education |  |  |  |  |
| No education | Ref. | Ref. | Ref. | Ref. |
| Primary | 0.8 (0.4–1.6) | 0.7 (0.5–1.1) | 0.6(0.5–0.9)** | 0.6(0.5–0.8)** |
| Secondary or above | 0.8 (0.3–1.8) | 0.8 (0.5–1.3) | 0.6(0.4–0.8)*** | **0.6(0.4–0.8)\*\*\*** |
| Partner's Alcohol Use |  |  |  |  |
| No | Ref. | Ref. | Ref. | Ref. |
| Yes | **3.5(1.1–11.4)\*** | **5.6(2.8–11.5)\*\*\*** | **3.1(2.2–4.4)\*\*\*** | **3.0(2.1–4.1)\*\*\*** |
| **Household Characteristics** |  |  |  |  |
| Wealth Index |  |  |  |  |
| Poor | Ref. | Ref. | Ref. | Ref. |
| Middle | 0.7 (0.4–1.4) | 0.6(0.4–1.0)* | 0.7(0.6–1.0)* | 0.7(0.6–0.9)* |
| Rich | 0.6 (0.2–1.3) | 0.4(0.3–0.7)*** | 0.7* (0.5–0.9) | **0.6 (0.5–0.8)\*\*\*** |
| Place of Residence |  |  |  |  |
| Urban | Ref. | Ref. | Ref. | Ref. |
| Rural | 0.7 (0.4–1.4) | 1.0 (0.7–1.5) | 1.1 (0.8–1.4) | 1.1 (0.8–1.4) |

**Noted:** Survey weights are applied to obtain weighted percentages: \* p < 0.05, \*\* p < 0.01, \*\*\* p < 0.001; **AOR** = adjusted odds ratio; **CI** = confidence interval; and **Ref.**= Reference.

Smartphone ownership is associated with reduced odds of both emotional violence (adjusted odds ratio [AOR] = 0.7; 95% CI (0.5–0.9) and IPV (AOR = 0.7; 95% CI (0.5–1.0) when compared to women without mobile phones. In contrast, low-frequency internet use correlates with elevated odds of experiencing emotional violence (AOR = 1.7; 95% CI (1.1–2.7) and IPV (AOR = 1.6; 95% CI (1.1–2.5) relative to women who do not use the internet.

Furthermore, women’s educational attainment appears to offer protection, as those with secondary or higher education exhibit lower odds of physical violence (AOR = 0.5, 95% CI (0.3– 0.9) and emotional violence (AOR = 0.7, 95% CI (0.5–1.0) compared to women without formal education. The educational level of the woman’s partner is also influential; women whose partners attained formal education (primary, secondary or higher) show reduced odds of emotional violence (AOR = 0.6, 95% CI (0.5–0.9); AOR = 0.6, 95% CI (0.4–0.8), respectively) and any IPV (AOR = 0.6, 95% CI (0.5–0.8) for both) compared to those whose partners have no education.

A strong positive association was observed between the partner’s alcohol use and violence types: sexual (AOR = 3.5; 95% CI (1.1–11.4), physical (AOR = 5.6; 95% CI (2.8–11.5), emotional (AOR = 3.1; 95% CI (2.2–4.4), and any IPV (AOR = 3.0; 95% CI (2.1–4.1).

Household wealth was inversely associated with IPV. Women in rich households had lower odds of physical, emotional, and IPV compared to those in the poorest households. Specifically, women in middle-wealth households had significantly lower odds of physical (AOR = 0.4; 95% CI (0.3–0.7) and IPV (AOR = 0.6; 95% CI (0.5–0.8).

In this adjusted model, motorcycle ownership and media exposure did not show statistically significant associations with IPV. Similarly, the association between women’s age and partner’s age with IPV was not statistically significant after adjusting for other factors. Place of residence (rural vs. urban) also did not emerge as an important predictor of IPV in this analysis.

### Interaction analyses on the factors associated with violence types

**Figure 1** presents the predicted probabilities of experiencing different forms of domestic violence (sexual, physical, emotional, and intimate partner violence [IPV]) as a function of the interaction between women’s mobile phone ownership status, partner’s alcohol consumption, and household wealth index. The data reveal a consistent pattern of elevated predicted probabilities for all violence types when partners consumed alcohol, irrespective of women’s mobile phone ownership or household wealth index. This finding aligns with extensive literature demonstrating the robust association between partner alcohol use and increased risk of IPV in Cambodia (8, 9, 27). In contrast, an inverse relationship is observed between household wealth index and the predicted probability of violence, with women in wealthier households demonstrating lower predicted probabilities across all categories of violence and partner alcohol consumption. This protective effect of economic resources has been documented in various contexts, suggesting that economic empowerment can reduce women’s vulnerability to violence (28, 29). The association between mobile phone ownership and predicted probabilities is more nuanced. While women with smartphone ownership tend to exhibit lower predicted probabilities of sexual and physical violence in the absence of partner alcohol consumption, this association is less pronounced, or attenuated, when partners consume alcohol. The complex role of technology in IPV dynamics, where it can offer both protective and risk factors, is increasingly recognized (12, 14, 30). These findings underscore the complex interplay of socio-economic and behavioral factors in predicting the likelihood of domestic violence.

**Fig 1.**
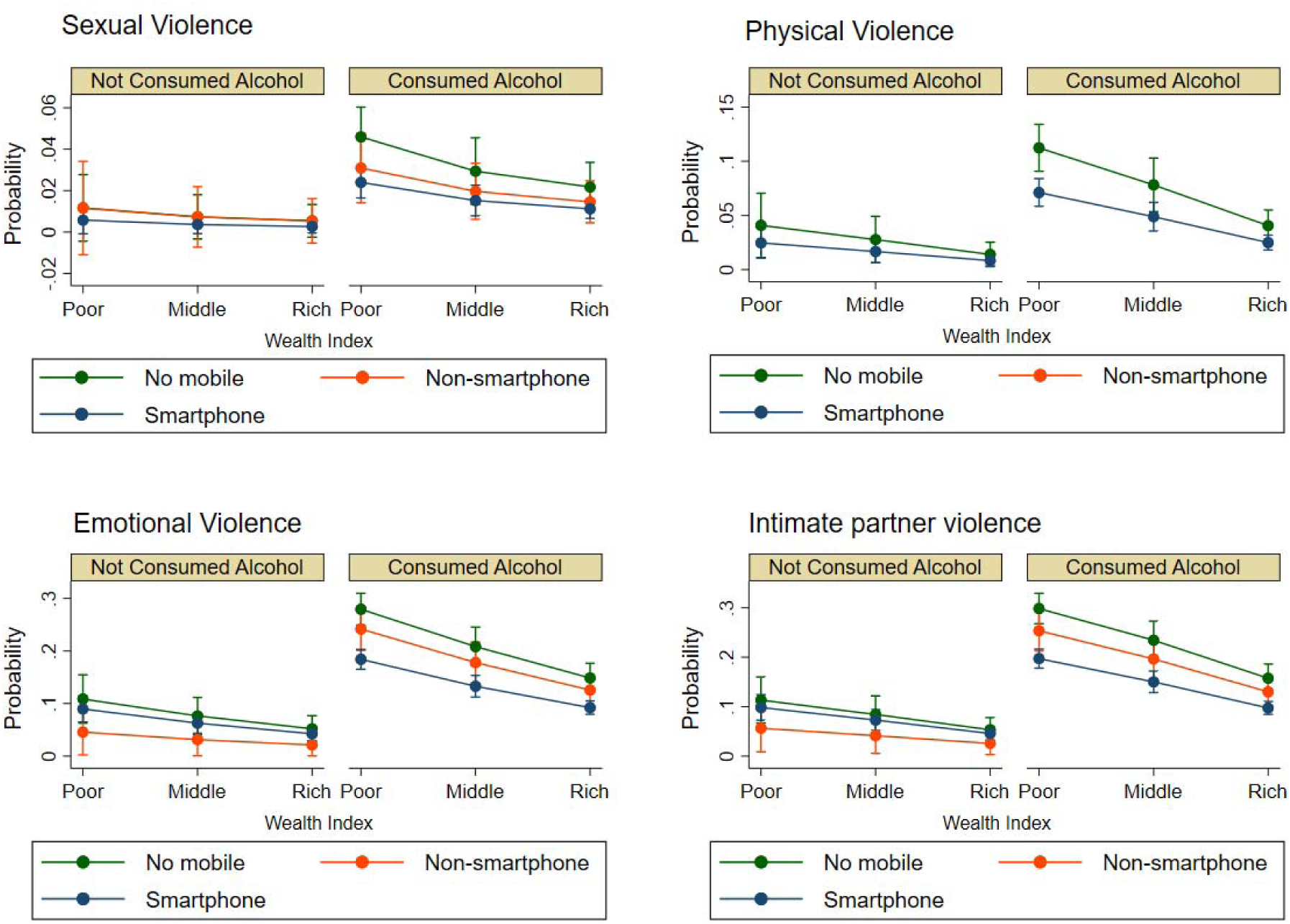
Predicted probability of domestic violence types. Illustrates the estimated probabilities resulting from the interaction between mobile phone ownership status, partner’s alcohol use, and household wealth index.

## Discussion

This study examined the relationship between digital access, mass media exposure, motorcycle ownership, and emotional, physical, intimate, sexual, and intimate partner violence (IPV) among Cambodian women aged 15–49, using recent data from the 2021–22 Cambodia Demographic and Health Survey (CDHS). This research contributes necessary evidence for low- and middle-income countries, particularly within Southeast Asia, where we found that 13.2% of Cambodian women reported experiencing IPV in the past year (emotional: 12.2%, physical: 4.4%, sexual: 1.9%), similar to these studies utilize CDHS 2021-2022 dataset (7–9). Furthermore, our analysis explored whether smartphone ownership and internet use act as protective or risk factors for IPV after adjusting for socioeconomic and behavioral covariates.

Our findings build upon the backdrop provided by recent 2021-2022 CDHS analyses. Shaikh (2025) highlighted the geographical clustering of IPV in areas with lower female autonomy and digital connectivity, underscoring the spatial dimension of IPV risk (31). Um et al. (2025) confirmed the strong association between high alcohol consumption and gender-based violence in Cambodia, advocating for integrated prevention programs (27). Banstola et al. (2025) demonstrated the influence of media exposure on sensitive health behaviors, suggesting the potential of information channels for positive change (15, 18). These studies identify digital access, spatial context, and substance use as critical leverage points for gender-sensitive interventions, issues that our analysis directly addresses at the individual level.

The protective association of smartphone ownership with a 30% reduction in the odds of any IPV after adjustment aligns with patterns observed in other South Asian contexts (32). This suggests that in Cambodia, smartphones may empower women by providing discreet access to support networks, vital information, and resources, thereby mitigating their vulnerability to violence. Conversely, the finding that infrequent internet use (less than daily) was associated with a 60% increase in the odds of emotional and overall IPV warrants further consideration. This may be indicative of a “backlash effect,” where limited and perhaps misunderstood internet engagement by women in a context of prevailing patriarchal norms can trigger suspicion, jealousy, and controlling behaviors from partners. Such intermittent use might be perceived as secretive or challenging to established traditional gender roles, leading to increased emotional abuse and overall IPV The lack of independent associations between mass media exposure and motorcycle ownership with IPV in our adjusted models suggests that mere access or exposure to these resources may not be sufficient to alter the underlying power dynamics that contribute to violence. In the case of media, the content of the messaging, rather than the frequency of exposure, may be the critical factor, as highlighted by Banstola et al.’s findings on reproductive health decisions (18, 21). The current media landscape in Cambodia may lack consistent and impactful IPV prevention content. Similarly, the absence of a significant association between motorcycle ownership and IPV indicates that increased physical mobility alone, without concomitant shifts in gender norms and women’s autonomy within the household, may not translate to a reduction in violence.

Consistent with Um et al.’s (2025) findings, partner alcohol consumption remained the most powerful predictor of IPV across all outcomes (adjusted OR ≈ 4–6) (9, 27). This reinforces the critical role of alcohol in escalating violence, emotional abuse, and controlling behaviors within intimate relationships in Cambodia. Furthermore, lower educational attainment and lower household wealth also emerged as significant predictors of IPV, underscoring the importance of addressing structural inequalities in violence prevention efforts.

Our findings suggest a dual digital strategy for IPV prevention in Cambodia. First, efforts should focus on expanding women’s private and secure access to smartphones and digital literacy training to maximize their safe and effective use. Second, there is a critical need to embed targeted anti-violence messaging and information about support services within popular digital platforms and mass media channels, emphasizing on the content of these communications. Concurrently, alcohol-harm reduction initiatives remain paramount. Given Shaikh’s (2025) use of spatial epidemiology to identify of IPV hot spots reas, technology-based interventions and awareness campaigns should be prioritized in these high-prevalence areas (31). Future research employing longitudinal designs and mixed-methods approaches is essential to understand better the dynamic interplay between digital empowerment, evolving gender norms, and IPV risk in Cambodia’s rapidly transforming information ecosystem.

### Strengths and limitations

This study benefits from several key strengths, including the use of a large, nationally representative sample from the 2021-22 CDHS, the application of validated and standardized DHS measures for assessing IPV, and the use of survey-weighted statistical methods, including multivariable logistic regression, to account for the complex survey design and control for critical sociodemographic covariates.

Despite these strengths, several limitations warrant consideration. The cross-sectional nature of the data precludes establishing causal relationships between the examined factors and IPV. Reporting bias, inherent in studies addressing sensitive topics like IPV, may also influence the findings. Furthermore, the CDHS dataset lacks specific measures of digital autonomy, such as whether women have private access to their devices or are subject to partner control. The media exposure variable’s focus on frequency rather than content represents another limitation. Finally, the study’s findings are specific to ever-partnered women and may not be generalizable to unmarried or non-cohabiting individuals.

### Conclusion

This study reveals a complex relationship between digital access and IPV risk among Cambodian women. While smartphone ownership offers a protective effect, potentially by enhancing access to support and resources, infrequent internet use is associated with increased risk, possibly due to triggering partner suspicion in contexts of restrictive gender norms. The lack of a strong independent association between media exposure and motorcycle ownership underscores the importance of content and normative change alongside access. Ultimately, partner alcohol use remains a critical and potent risk factor, highlighting the persistent need to address both behavioral and structural determinants, including poverty and low education, in IPV prevention efforts. To effectively reduce IPV, programs should prioritize expanding safe and private digital access for women, integrating digital literacy and secure technology use training. Public messaging through diverse platforms should actively promote respectful relationships, positive masculinity, gender equality, focusing on impactful content. Simultaneously, community-level alcohol-reduction programs and poverty-alleviation initiatives, such as cash support and job skills training, are crucial for addressing the underlying structural factors contributing to women’s vulnerability to IPV. Future research should employ longitudinal designs and qualitative methods to elucidate further the dynamic interplay between digital access, partner dynamics, and women’s safety in Cambodia.

## Data Availability

The Cambodia Demographic and Health Survey data are publicly available from the website: (URL: https://www.dhsprogram.com/data/dataset_admin).

## Acknowledgments

We thank the Ministry of Planning and the Ministry of Health of Cambodia for granting access to the CDHS dataset. We also acknowledge ICF International for technical support in survey implementation.

## Supporting information

**S1 Table.** Variance Inflation Factor (VIF) for Sexual Violence Model

**S2 Table.** Variance Inflation Factor (VIF) for Physical Violence Model

**S3 Table**. Variance Inflation Factor (VIF) for Emotional Violence Model

**S4 Table**. Variance Inflation Factor (VIF) for Any Intimate Partner Violence Model

